# Efficacy and Safety of Ensartinib in the Treatment of Non-Small Cell Lung Cancer: A Systematic Review of Clinical Trials

**DOI:** 10.1101/2025.06.02.25328724

**Authors:** Trushdeep Agrawal

## Abstract

Anaplastic lymphoma kinase (ALK) rearrangements are the therapeutic targets in non-small cell lung cancer (NSCLC). Ensartinib, a second-generation ALK tyrosine kinase inhibitor (TKI), was recently approved by the FDA for the treatment of ALK-positive NSCLC. In this systematic review, we identified five studies including a total of 621 participants, by searching PubMed, Scopus, and Embase through May 2025. The phase I clinical trials suggested a recommended phase II dose (RP2D) of 225 mg. Ensartinib demonstrated favourable efficacy across dose-escalation, phase II and phase III trials. In treating naïve patients, ORRs ranged from 80–81%, with median PFS reaching up to 26.2 months. In pre-treated cases, efficacy was also notable, including intracranial response upto 70%. Phase III trial confirmed superior PFS with ensartinib compared to crizotinib. Common AEs include rash, transaminase elevations, and gastrointestinal symptoms, which were mostly manageable and grade 1–2 in severity. Ensartinib is a highly effective and tolerable option for ALK-positive NSCLC. Further studies are needed to assess long-term outcomes and to optimize its use in a molecularly diverse patient population.

## Introduction

Lung cancer is the leading cause of cancer deaths and Non-small cell lung carcinoma (NSCLC) is the most frequent subtype of lung cancer. Tyrosine kinase inhibitors and immunotherapy has led to improved survival benefits in selected patients. However, in patients suffering from NSCLC, the overall cure and survival rates remain low, especially in case of metastatic disease. Therefore, continued research into new drugs is required to expand the clinical benefits and improve overall prognosis [1, 2].

Anaplastic lymphoma kinase (ALK) is a proto-oncogene which encodes anaplastic lymphoma kinase. In 2007, ALK rearrangements were first identified in NSCLC. ALK+ NSCLC patients are generally younger, with no smoking history, and have adenocarcinoma as the most common histological subtype [3, 4, 5].

Because of the gene fusion-driven nature of ALK+ NSCLC, tyrosine kinase inhibitors (TKIs) have been developed to treat this unique disease [6]. On December 18, 2024, the Food and Drug Administration approved ensartinib for adult patients with ALK+ NSCLC. Ensartinib is a novel second-generation ALK-TKI which improves the activity on CNS metastasis [7].

Ensartinib was developed to overcome the limitations of prior generations of TKIs and improve the overall prognosis and the intracranial activity. Ensartinib was found to be ten times more potent than crizotinib at inhibiting the growth of ALK+ lung cancer cell lines [8].

This study focuses on the clinical outcomes of trials conducted in NSCLC patients. Various studies across the various countries have been included and a comprehensive understanding of the current clinical trial scenario for ensartinib has been provided in the study. The objective of this study is to evaluate the clinical outcomes and safety of ensartinib in NSCLC patients.

## Methodology

This systematic review was conducted in accordance with the PRISMA 2020 guidelines [9] and the recommendations outlined in the Cochrane Handbook for Systematic Reviews [10].

### Search strategy

We performed a comprehensive search of PubMed, Scopus, and Embase, from their respective inceptions to May 2025. Our search terms targeted NSCLC and ensartinib, using a combination of keywords and subject headings such as (“Ensartinib” OR “X-396” OR “Ensartinib”) AND (“non-small cell lung cancer” OR “NSCLC” OR “non small cell lung carcinoma” OR “Lung Neoplasms”) AND (“clinical trial” OR “clinical study” OR “phase 1” OR “phase 2” OR “phase 3”). Full data can be found in supplementary data table 1.

**Table 1.** Baseline characteristics for patients in the trials.

| Author, Year | Drug | Sample size | Age(years) | Male n (%) | Female n (%) | ECOG performance status n (%)<br>0/ 1/ 2 |
| --- | --- | --- | --- | --- | --- | --- |
| Horn et al., 2018 (NCT01625234) | Ensartinib<br>1. Dose escalating (25- 250 mg)<br>2. Dose expansion (225 mg) | 97<br>1. 37<br>2. 60 | 56 (median)<br>21- 83<br>(range) | 48 (49) | 49 (51) | 0: 31 (32),<br>1: 66 (68),<br>2: - |
| Yang et al., 2019 (NCT03215693) | Ensartinib (225 mg) | 156 | 52 (median)<br>44- 60<br>(range) | 81 (52) | 75 (48) | 0: 25 (16)<br>1: 128 (82)<br>2: 3 (2) |
| Horn et al., 2021 (eXalt3) (NCT02767804) | 1. Ensartinib (225 mg)<br>2. Crizotinib (250 mg) | Total: 290 (randomized 1:1)<br>Ensartinib (n= 143)<br>Crizotinib (n= 147) | 54 (median)<br>25- 90<br>(range) | 149 (51.4) | 141 (48.6) | Ensartinib<br>0 or 1: 136 (95.1)<br>2: 7 (4.9)<br>Crizotinib<br>0 or 1: 139/146 (95.2)<br>2: 7/146 (4.8) |
| Ma et al., 2022 (NCT02959619) | Ensartinib<br>1. Dose escalation (150, 200, 225, and 250 mg)<br>2. Dose expansion (225 mg) | 48<br>1. 16<br>2. 32 | 48 (median)<br>23- 77<br>(range) | 23 (47.9) | 25 (52.1) | 0: 21 (43.8)<br>1: 27 (56.2)<br>2: - |
| Xia et al., 2025 (ChiCTR2100048767) | Ensartinib (225 mg) | 30 | 69 (median)<br>47- 83<br>(range) | 14 (46.7) | 16 (53.3) | 0: -<br>1: 24 (80)<br>≥2: 6 (20) |

### Eligibility criteria

We applied the following criteria based on the PICO framework:

- Population (P): Adult patients with NSCLC
- Intervention (I): Ensartinib used as monotherapy or in combination with other drugs, at any dosage or schedule.
- Comparison (C): No formal comparator was required.
- Outcomes (O): Measures of response to treatment and safety

Clinical trials that reported relevant efficacy and safety outcomes were included. Studies were excluded if they were observational studies, brief reports, protocols, or conference abstracts.

### Selection of studies

Titles and abstracts were screened using Rayyan software after removing duplicates. Full-text articles were then evaluated. Only studies meeting the inclusion criteria were included in the final analysis.

### Data synthesis and outcomes

Data from each included study was extracted. The “Summary sheet” contains the key study details such as author, country, design/phase, total sample size, interventional details, and reported efficacy and safety outcomes. A separate “Baseline Sheet” recorded demographic and clinical characteristics such as age, sex, and Eastern Cooperative Oncology Group (ECOG) performance status. The “Outcome sheet” summarized sample sizes, clinical outcomes and safety of the participants receiving the drug.

### Quality assessment

The quality of each trial was assessed using the Risk of Bias In Non-randomized Studies of Intervention (ROBINS-I) focusing on the following seven domains. Each domain was assessed if it was (Low risk), (Moderate risk), or (Serious risk) or (Critical risk) of bias.

## Results Search results

The initial database search yielded a total of 65 articles from PubMed, Scopus, and Embase. After removing 10 duplicates, 55 articles remained for title and abstract screening. We then evaluated 38 articles of which 33 were excluded for various reasons (see Figure 1 for details). 5 studies were found to be eligible for the study and were included in the systematic review. A PRISMA Flow diagram illustrating the selection process is presented in Figure 1.

**Figure 1.**
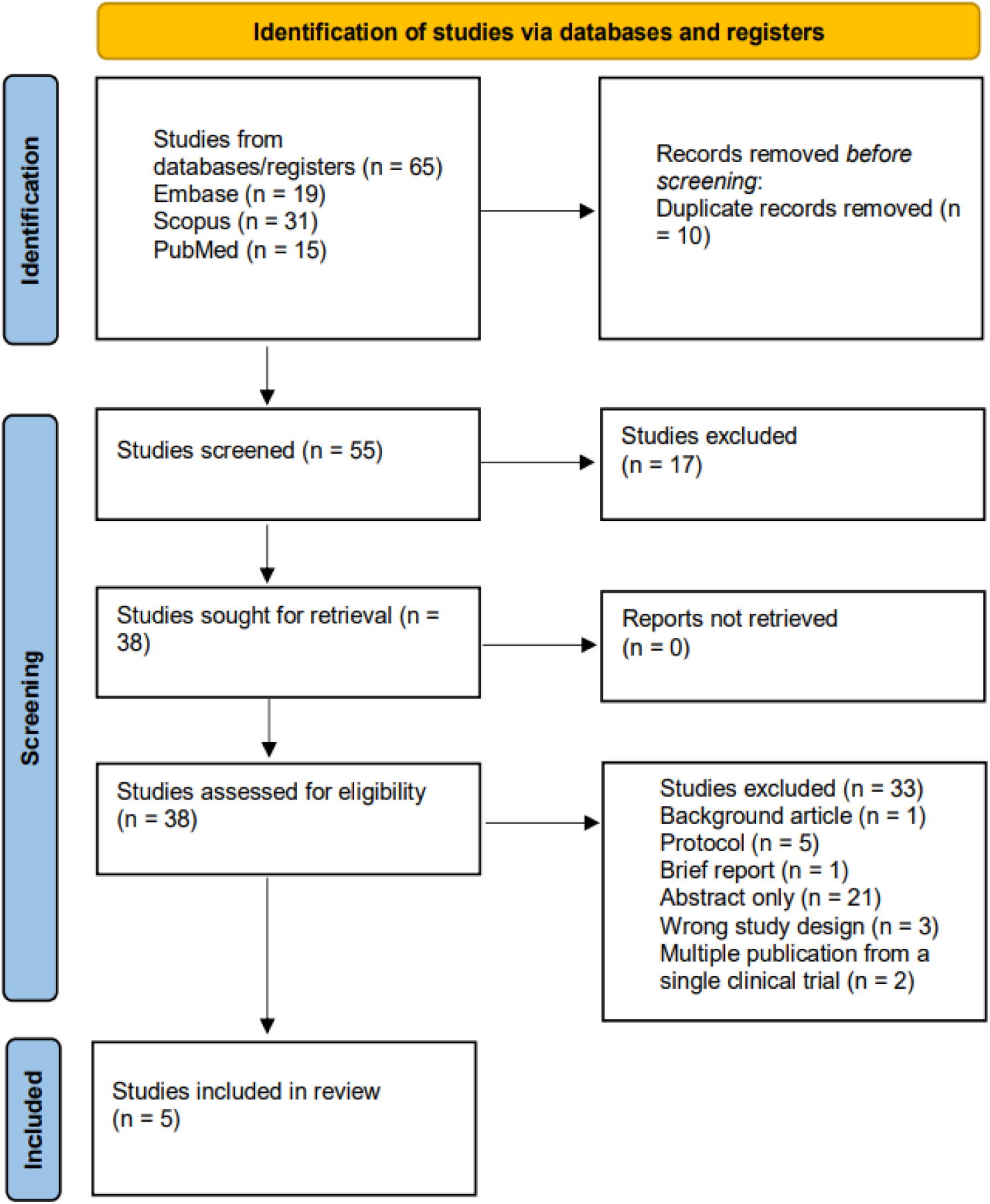
PRISMA Flowchart for this systematic review.

### Baseline characteristics of patients

Table 1 shows the baseline characteristics of participants across the five studies included, encompassing a total of 621 participants. The median age in the individual studies ranged from 48 to 69 years. All of the studies reported a nearly equal proportion of both the sexes. In terms of functional status, the majority of the patients had an ECOG performance status of 0 to 1, indicating they were generally able to carry out most of their daily activities.

### Studies and intervention characteristics

All five included studies were open-label, multi-center trials conducted in multiple countries. Majority of these studies were conducted in the United States and China. These trials evaluated the safety and efficacy of ensartinib, in patients with NSCLC. Two of these studies performed a dose-escalation and dose-expansion trial. Most of the trials were performed at a dose/regimen of 225 mg once a day (QD). One of these studies was a phase III randomized clinical trial, with the comparator crizotinib, with a dose/regimen of 250 mg twice a day (BD). Detailed intervention characteristics are summarized in Table 2.

**Table 2.**
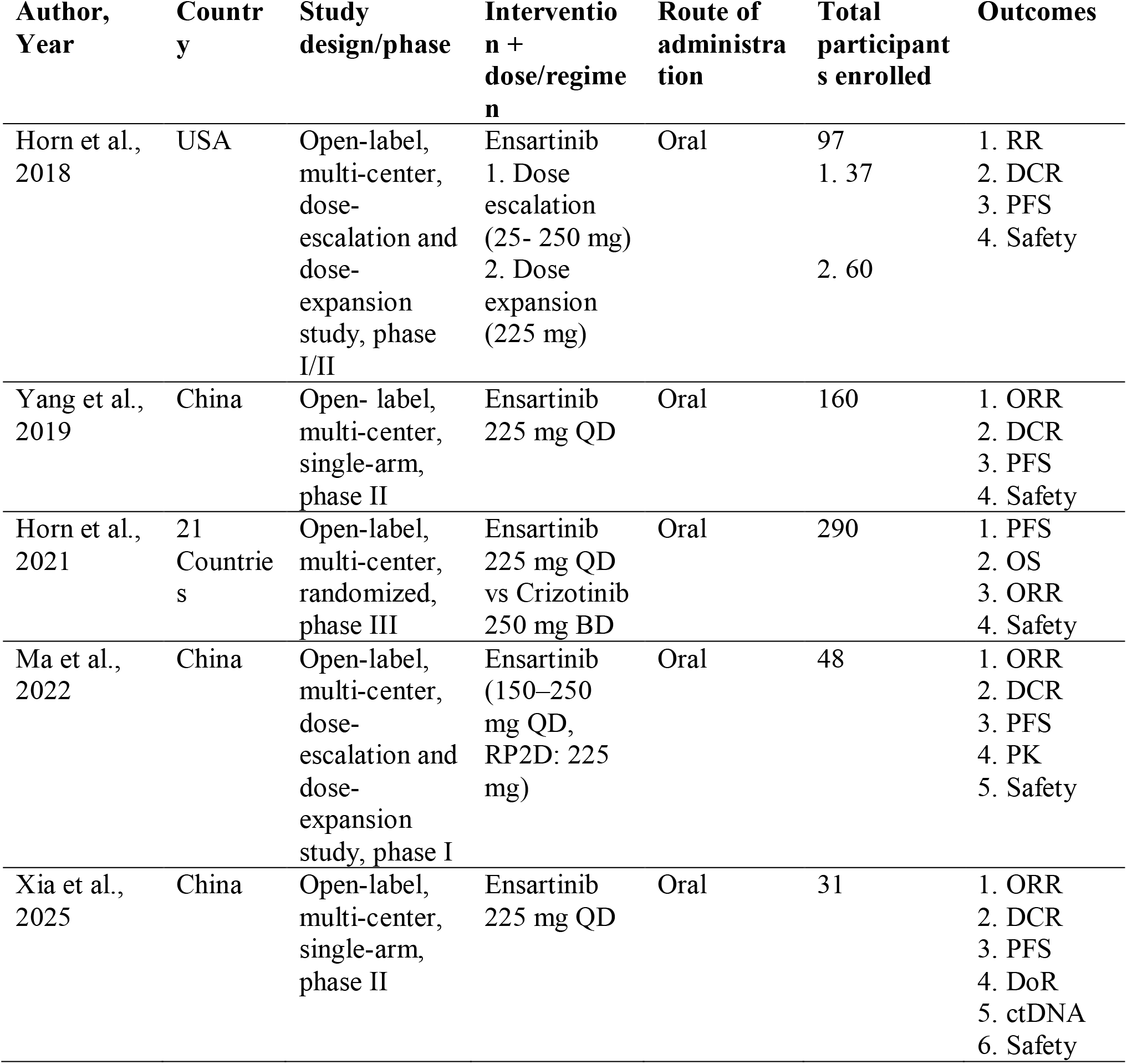
Summary table for the included trials.

| Author, Year | Country | Study design/phase | Intervention + dose/regimen | Route of administration | Total participants enrolled | Outcomes |
| --- | --- | --- | --- | --- | --- | --- |
| Horn et al., 2018 | USA | Open-label, multi-center, dose-escalation and dose-expansion study, phase I/II | Ensartinib<br>1. Dose escalation (25- 250 mg)<br>2. Dose expansion (225 mg) | Oral | 97<br>1. 37<br>2. 60 | 1. RR<br>2. DCR<br>3. PFS<br>4. Safety |
| Yang et al., 2019 | China | Open-label, multi-center, single-arm, phase II | Ensartinib 225 mg QD | Oral | 160 | 1. ORR<br>2. DCR<br>3. PFS<br>4. Safety |
| Horn et al., 2021 | 21 Countries | Open-label, multi-center, randomized, phase III | Ensartinib 225 mg QD vs Crizotinib 250 mg BD | Oral | 290 | 1. PFS<br>2. OS<br>3. ORR<br>4. Safety |
| Ma et al., 2022 | China | Open-label, multi-center, dose-escalation and dose-expansion study, phase I | Ensartinib (150–250 mg QD, RP2D: 225 mg) | Oral | 48 | 1. ORR<br>2. DCR<br>3. PFS<br>4. PK<br>5. Safety |
| Xia et al., 2025 | China | Open-label, multi-center, single-arm, phase II | Ensartinib 225 mg QD | Oral | 31 | 1. ORR<br>2. DCR<br>3. PFS<br>4. DoR<br>5. ctDNA<br>6. Safety |

### Quality assessment

The ROBIN-I tool was used to assess risk the quality of studies included. Of the five studies included, four studies were deemed low risk of bias and one was deemed moderate risk of bias (Table 3).

**Table 3.**
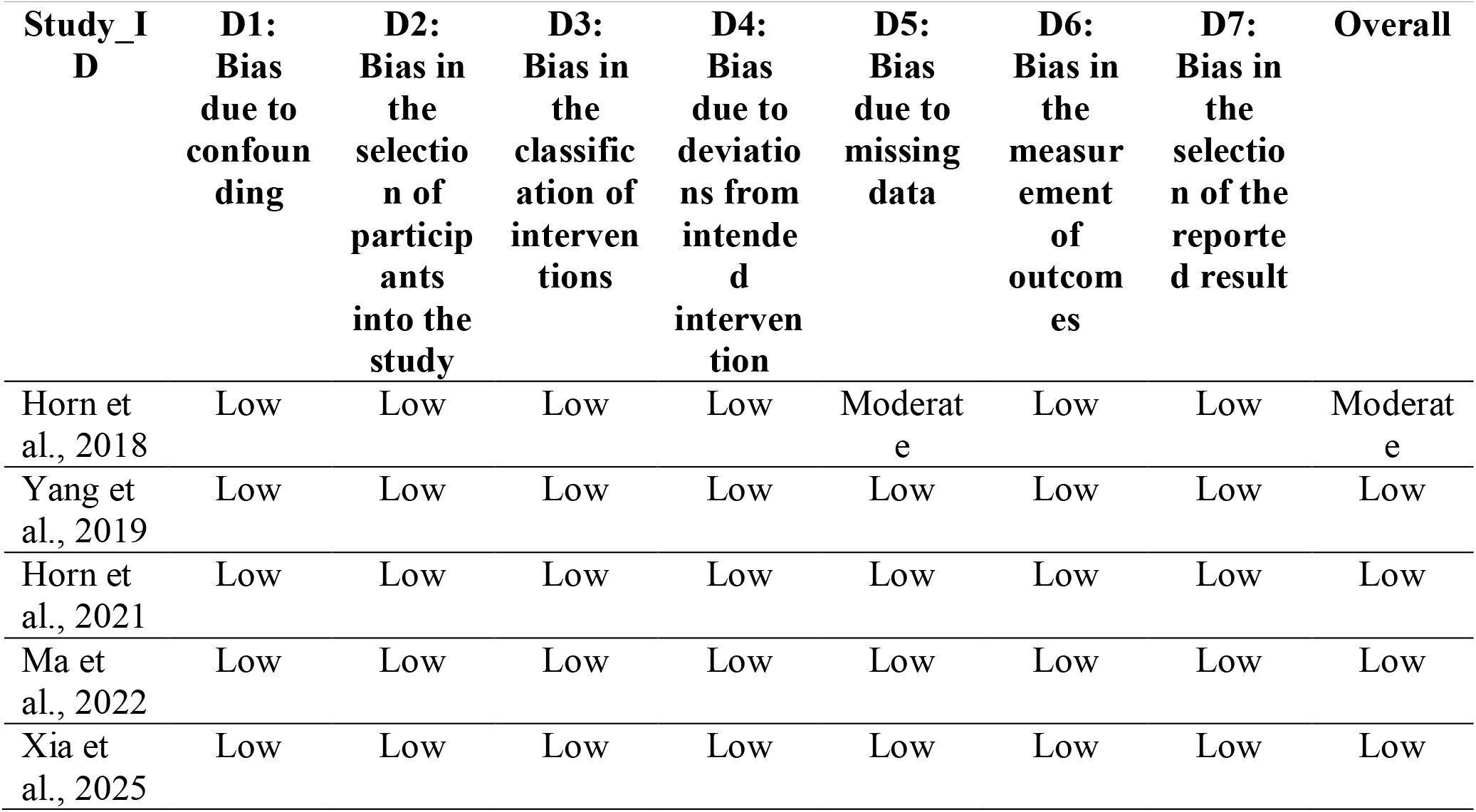
ROBIN-I for included studies.

| **Study_I**<br>**D** | **D1:**<br>**Bias**<br>**due to**<br>**confoun**<br>**ding** | **D2:**<br>**Bias in**<br>**the**<br>**selectio**<br>**n of**<br>**particip**<br>**ants**<br>**into the**<br>**study** | **D3:**<br>**Bias in**<br>**the**<br>**classific**<br>**ation of**<br>**interven**<br>**tions** | **D4:**<br>**Bias**<br>**due to**<br>**deviatio**<br>**ns from**<br>**intende**<br>**d**<br>**interven**<br>**tion** | **D5:**<br>**Bias**<br>**due to**<br>**missing**<br>**data** | **D6:**<br>**Bias in**<br>**the**<br>**measur**<br>**ement**<br>**of**<br>**outcom**<br>**es** | **D7:**<br>**Bias in**<br>**the**<br>**selectio**<br>**n of the**<br>**reporte**<br>**d result** | **Overall** |
| --- | --- | --- | --- | --- | --- | --- | --- | --- |
| Horn et al., 2018 | Low | Low | Low | Low | Moderate | Low | Low | Moderate |
| Yang et al., 2019 | Low | Low | Low | Low | Low | Low | Low | Low |
| Horn et al., 2021 | Low | Low | Low | Low | Low | Low | Low | Low |
| Ma et al., 2022 | Low | Low | Low | Low | Low | Low | Low | Low |
| Xia et al., 2025 | Low | Low | Low | Low | Low | Low | Low | Low |

## Efficacy and safety outcomes

### Dose-escalation and dose-expansion study

Two studies, Horn et al. 2018 [11] and Ma et al. 2021 [12], studied the efficacy and safety of ensartinib in a total of 145 participants (97 patients in Horn et al. 2018 and 48 patients in Ma et al. 2021).

#### Horn et al. 2018 [11]

This study was the first-in-human phase I/II trial of ensartinib. A total of 97 participants were included, with 37 in the dose-escalation phase and 60 in the dose-expansion phase. Among these individuals, 82% had undergone at least one prior systemic treatment for advanced disease. Four participants had head and neck cancer, two had colorectal cancer, and one had both non-small cell lung cancer and breast cancer. Throughout the trial, the maximum tolerated dose was not established, leading to the recommended phase II dose (RP2D) being set at 225 mg due to the rash occurrences noted at 250 mg.

In the 60 ALK-positive efficacy evaluable cohort treated with doses of 200 mg or more, the response rate (RR) was 60% (95% confidence interval [CI], 47.4−71.4) with a median progression-free survival (mPFS) of 9.2 months (95% CI, 5.6−11.7). Thirteen patients achieved stable disease (SD) after at least 2 treatment cycles, resulting in a disease control rate (DCR) of 81.7%, while 11 exhibited progressive disease (PD) as their best response. Among the 15 ALK TKI-naïve patients, 12 had a partial response (PR), yielding an RR of 80% (95% CI, 54.8−93.0) and a mPFS of 26.2 months. In patients who had only been treated with crizotinib before the trial, the RR was 69% (95% CI, 50.8−82.7) with a mPFS of 9 months (95% CI, 5.6−11.7). For the 14 patients with baseline CNS target lesions, 2 had a complete cranial response, and seven attained an intracranial partial response, resulting in an RR of 64.3% (95% CI, 38.8−83.7).

Adverse events (AEs) related to treatment were observed in 86% of the participants. The majority of these were classified as Grade 1-2, while 22 out of 97 individuals (23%) experienced Grade 3-4 AEs. Two subjects encountered dose-limiting toxicity (DLTs), which were resolved after pausing the treatment and resuming it at a lower dose. The most frequently reported AEs included rash (56%), nausea (36%), pruritus (28%), vomiting (26%), and fatigue (22%). It was observed that when ensartinib was taken with food, the frequency and severity of nausea and vomiting were reduced.

#### Ma et al. 2022 [12]

In this trial, a total of 48 participants were enrolled, 37 (77.1%) of these participants were ALK TKI-naïve, 11 (22.9%) of these participants had previously received TKIs like crizotinib, certinib or alectinib. The objective response rate (ORR) for all patients was 64.6%, the disease control rate (DCR) was 81.3% and the mPFS was 16.79 months. Among TKI-naïve patients ORR was 81.3% and mPFS was 25.73 months. Among treated ALK+ patients, ORR was 45.5% and mPFS was 4.14 months. For patients with brain metastasis, the overall response rate (ORR) was 66.7%, and the median progression-free survival (mPFS) was 22.90 months. Of all the treatment-related adverse events (AEs), 27% (13 out of 48) were classified as Grade 3-5. The majority of AEs were Grade 1-2. Serious adverse events were reported in 10 patients, accounting for 20.8%. The most frequently occurring AEs included rash (87.5%), elevation of transaminases (60.4%), pruritus (45.8%), and increased creatinine levels (35.4%). Two dose-limiting toxicities (DLTs) were noted in the cohort receiving the 250 mg dose, leading to a recommended phase 2 dose (RP2D) of 225 mg.

### Phase II trials

Two trials, Yang et al., 2019 [13] and Xia et al., 2025 [14] were conducted as open-label, multi-centre, single-arm, phase II trials in China. A total of 191 participants were enrolled in these studies (160 participants in Yang et al., 2019 and 31 participants in Xia et al., 2025).

#### Yang et al., 2019 [13]

About160 patients participated in the study, having received at least one dose of ensartinib. Four were later excluded due to violations of the inclusion criteria. Around 137 (88%) of the patients transitioned from crizotinib to ensartinib. Among the 147 patients whose responses were evaluable by the review committee, 76 (52%) showed a confirmed partial response, 61 (41%) had stable disease, 10 (7%) experienced disease progression, resulting in an objective response rate of 52% (95% CI 43–60) and a disease control rate of 93% (95% CI 88–97). The time taken to achieve the first response among participants was recorded as 1.3 months. The median progression-free survival in the full set of analysis was found out to be 9·6 months (95% CI, 7·4–11·6). Among 97 patients with baseline brain metastasis, 40 (41%) patients showed responses that could be assessed by the review committee. An objective intracranial response was noted in 28 patients (70% [95% CI 53–83]) and intracranial disease control in 39 patients (98% [87–100]).

At least one treatment-related adverse event was seen among 145 (91%) of 160 patients, which were mostly grade 1 or 2. The most common AEs were rash (89 [56%]), increased alanine aminotransferase concentrations (74 [46%]), and increased aspartate aminotransferase concentrations (65 [41%]).

Later, another couple of studies were published from the same clinical trial NCT03215693, which have not been included in this review [16, 17].

#### Xia et al., 2025 [14]

A total of 31 patients were enrolled. As a result of irregular follow-up, one patient was excluded from the trial. All these patients carried METex14. Out of the total, 5 patients (16.67%) were identified with brain metastases at the beginning of the study. The median duration of follow-up was noted to be 9.2 months, with an overall response rate (ORR) of 53.3% (16/30; 95% CI, 35.5– 71.2) and a disease control rate (DCR) of 86.7% (26/30; 95% CI, 74.5–98.8), the tumor shrinkage rate was found to be 33% (95% CI 24.7–41.4), the median time to response was found to be 0.93 months (95% CI 0.83–1.13). The median progression-free survival (mPFS) was determined to be 6.0 months (95% CI, 3.0–8.8). The median duration of response (mDoR) was recorded at 7.9 months (95% CI, 4.8–8.7). The median overall survival (mOS) was determined to be 11.8 months (95% confidence interval, 7.1–16.5). Within the group of 5 patients with baseline brain metastases, 4 (80%) exhibited partial responses, with an mPFS of 9.5 months (95% CI 2.2–9.6) and a DoR of 8.5 months (95% CI 1.2–8.7). This study indicated a significantly lower overall response rate (ORR) of 33.3% in patients with concurrent TP53 mutations, compared to an ORR of 61.9% among those with wild-type TP53. Adverse events (AEs) were documented in 24 patients (80%), with 7 (23.3%) classified as Grade 3. No Grade 4 or 5 AEs were observed. The most frequently reported AEs included rash (46.7%), followed by anemia (23.3%), elevated ALT (23.3%), elevated AST (23.3%), and pruritus (20%). There were no serious AEs or treatment-related fatalities reported.

### Phase III randomized clinical trial

#### Horn et al., 2021 [15]

This was a phase III clinical trial that was open-label and multi-centered. This research included 290 participants who were randomly assigned (1:1) to receive either ensartinib at a daily dose of 225 mg or crizotinib at a dose of 250 mg taken twice a day. The research demonstrated that the median progression-free survival (mPFS) in the intent-to-treat (ITT) group was greater in those receiving ensartinib compared to those on crizotinib, with durations of 25.8 months (range, 0.03-44.0 months) against 12.7 months (range, 0.03-38.6 months) respectively. The hazard ratio was found to be 0.51 ([95% CI, 0.35-0.72]; log-rank P < .001). The median follow-up duration for patients treated with ensartinib was 23.8 months (with a range from 0 to 44 months), while it was months (with a range from 0 to 38 months) for those on crizotinib. In the modified intention-to-treat (mITT) population, the ensartinib group did not reach a median progression-free survival (PFS), in contrast, the crizotinib group had a median PFS of 12.7 months (95% confidence interval, 8.9-16.6 months). The hazard ratio in this case was 0.45 ([95% CI, 0.30-0.66]; log-rank P < .001). The intracranial response rate for patients taking ensartinib was noted to be 63.6%, in contrast to 21.1% for those on crizotinib with target brain metastases at the start. For patients who did not have brain metastases, the progression-free survival (PFS) for ensartinib was not established, while crizotinib exhibited a PFS of 16.6 months. There were only 3 instances of grade 4 adverse events (AEs) linked to ensartinib. The occurrence of serious treatment-related AEs was minimal for ensartinib, affecting 11 patients (7.7%), that primarily involving rash and liver toxicity. AEs that led to a dose reduction of ensartinib were noted in 34 out of 143 patients (23.8%), and dose was discontinued in 13 out of 143 patients (9.1%).

## Strengths and limitations

A major strengths of this review lies in its thorough assessment of ensartinib through various clinical stages, such as dose-escalation, dose-expansion, phase II, and randomized phase III trials. This tiered system facilitates a clearer vision of the drug’s efficacy and safety. By using information from the early-phase trials along with a large, randomized trial, this review provides a complete picture of clinical parameters like ORR, RR, mPFS, safety, and intracranial activity. The use of heterogeneous patient populations with CNS metastases and prior treatment statuses increases the applicability and relevance of the results. The uniform determination of a phase II dose recommendation between trials substantiates the reproducibility and translatable value of the data. Several limitations of this review, however, must be noted. A number of studies included were early-phase, single-arm trials without control groups.

## Conclusion

Ensartinib has emerged as a potent next-generation ALK inhibitor with prominent efficacy and tolerability across multiple trials. In both treatment-naïve and pretreated ALK-positive NSCLC patients. Ensartinib has shown robust ORR, mPFS, and encouraging intracranial activity, particularly in patients with brain metastases.

It has a manageable safety profile, with rash and transaminase elevations as most common AEs. Phase III clinical trial proves its superiority over crizotinib. The inconsistent responses among various molecular subgroups and the need for long-term outcome data shows the necessity for continued investigation. This review highlights the clinical promise of ensartinib while advocating for further research to refine its use in therapeutic oncology.

## Data Availability

All data produced in the present work are contained in the manuscript.

## Supplementary Data

**Supplementary Table 1.**
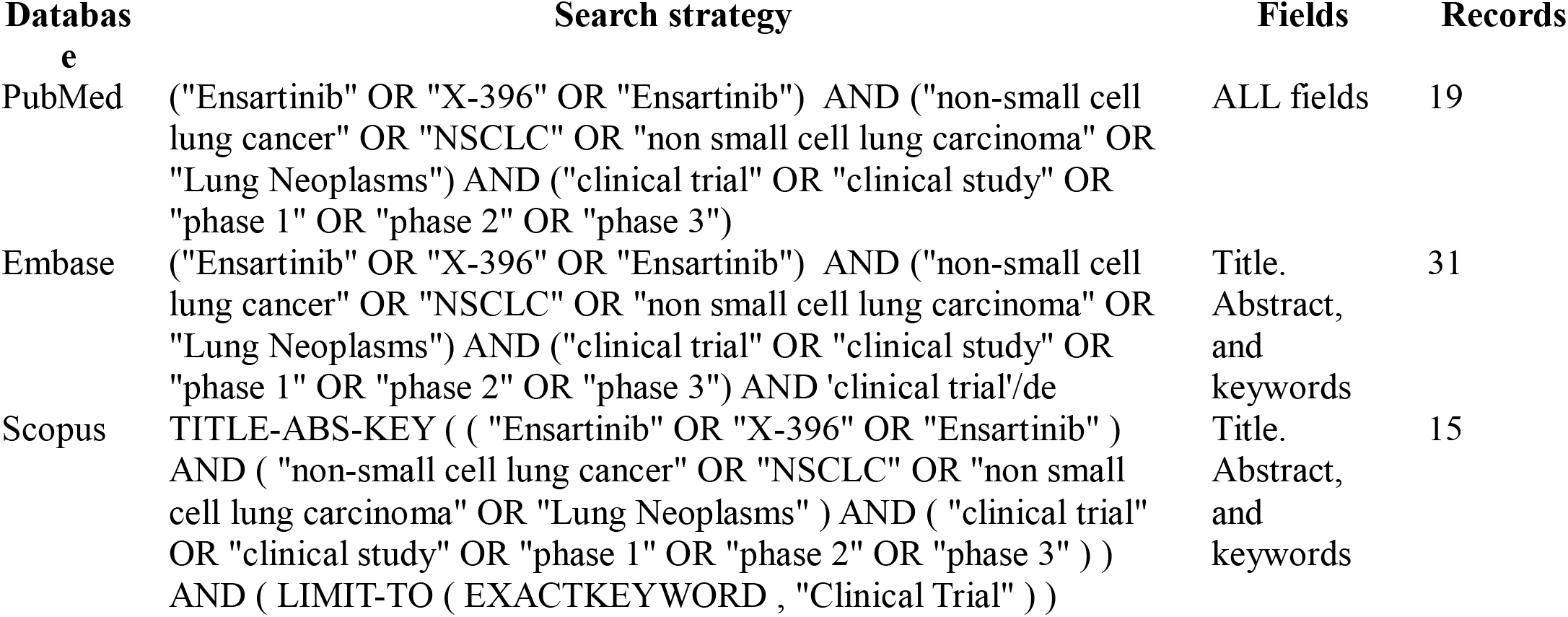
Search stratergy.

