## Supplemental Table 1 for "Efficacy and Safety of Ensartinib in the Treatment of Non-Small Cell Lung Cancer: A Systematic Review of Clinical Trials"

Trushdeep Agrawal^1,2^

^1^ Shri Vasantrao Naik Government Medical College, Yavatmal, Maharashtra, India

^2^ Sujan Surgical Cancer Hospital and Amravati Cancer Foundation, Amravati, Maharashtra, India

Corresponding Author:

Trushdeep Agrawal

| **Database** | **Search strategy** | **Fields** | **Records** |
| --- | --- | --- | --- |
| PubMed | ("Ensartinib" OR "X-396" OR "Ensartinib") AND ("non-small cell lung cancer" OR "NSCLC" OR "non small cell lung carcinoma" OR "Lung Neoplasms") AND ("clinical trial" OR "clinical study" OR "phase 1" OR "phase 2" OR "phase 3") | ALL fields | 19 |
| Embase | ("Ensartinib" OR "X-396" OR "Ensartinib") AND ("non-small cell lung cancer" OR "NSCLC" OR "non small cell lung carcinoma" OR "Lung Neoplasms") AND ("clinical trial" OR "clinical study" OR "phase 1" OR "phase 2" OR "phase 3") AND 'clinical trial'/de | Title. Abstract, and keywords | 31 |
| Scopus | TITLE-ABS-KEY ( ( "Ensartinib" OR "X-396" OR "Ensartinib" ) AND ( "non-small cell lung cancer" OR "NSCLC" OR "non small cell lung carcinoma" OR "Lung Neoplasms" ) AND ( "clinical trial" OR "clinical study" OR "phase 1" OR "phase 2" OR "phase 3" ) ) AND ( LIMIT-TO ( EXACTKEYWORD , "Clinical Trial" ) ) | Title. Abstract, and keywords | 15 |

Supplementary Table 1. Search stratergy
